# Novel type 1 diabetes polygenic scores improve identification of type 1 diabetes in diverse populations

**DOI:** 10.64898/2026.09.04.26361335

**Authors:** Stella Nam, Raymond J. Kreienkamp, Jiang Li, Ravi Mandla, Huy Tran, Katherine Taylor, Maheak Vora, Alicia Huerta-Chagoya, Jun Wei, Ashley J. Mulford, Alan R. Sanders, Jianfeng Xu, Liana K. Billings, Bogdan Pasaniuc, David J. Carey, Uyenlinh L. Mirshahi, Jose C. Florez, Alisa K. Manning, Josep M. Mercader, Miriam S. Udler, Aaron J. Deutsch

## Abstract

Most polygenic scores (PS) for type 1 diabetes were developed using European (EUR) ancestry datasets, limiting performance in non-European (non-EUR) populations. We evaluated novel type 1 diabetes PS across diverse populations to assess whether newer models improve prediction in underrepresented populations. Seven type 1 diabetes PS (T1D GRS2, T1D GRS2′, T1D GRS_HLA_, T1GRS, TA-PS, TA-PS (S), T1D MAPS) were evaluated in All of Us and Mass General Brigham Biobank. Predictive performance was assessed using the area under the receiver operating characteristic curve (AUC). T1D GRS2 and T1D MAPS showed the highest AUC in All of Us, while other scores demonstrated comparatively lower performance. These highest-performing scores were then evaluated in a meta-analysis across three additional biobanks (Genomic Health Initiative at Endeavor Health, Penn Medicine BioBank, Geisinger MyCode). Among 2,782 individuals with type 1 diabetes and 546,577 controls, T1D MAPS showed comparable performance to T1D GRS2 in EUR populations but significantly improved discrimination in non-EUR populations (meta-analysis ΔAUC=0.049, *p*=1.7×10^−7^). Overall, T1D MAPS improves prediction in non-EUR populations, highlighting the importance of multi-ancestry approaches for equitable genetic risk prediction. These findings could inform precision medicine in diverse populations.

---

Type 1 diabetes is a complex disease influenced by environmental and genetic factors, with heritability estimated over 50% [1]. Polygenic scores (PS), which integrate risk variants across the genome, are powerful tools to assess genetic risk for type 1 diabetes. PS have strong predictive performance in European (EUR) populations. However, their accuracy often declines in non-European (non-EUR) populations, due to differences in genetic architecture and underrepresentation of diverse ancestries in genome-wide association studies (GWAS) [2,3]. Recently, several novel type 1 diabetes PS were developed. In this study, we evaluated established and novel type 1 diabetes PS across ancestries and assessed whether newer approaches improve prediction among underrepresented populations.

## Methods

We evaluated several type 1 diabetes PS (T1D GRS2 [4], T1D GRS2′ [5], T1D GRS_HLA_ [6)], T1GRS [7], TA-PS [8], TA-PS (S) [8], T1D MAPS [9]) across two biobanks: All of Us (AoU) [10] and Mass General Brigham Biobank (MGBB) [11]. In AoU, we were unable to impute HLA variants on the Michigan Imputation Server due to privacy restrictions; therefore, T1GRS was evaluated only in MGBB. We did not re-evaluate an African-specific PS [12], which was assessed in a prior publication [9]. Genetic ancestry was classified as EUR or non-EUR using principal component analysis. Non-EUR participants were further stratified into African (AFR), Admixed American (AMR), and Other (OTH) ancestries. Due to privacy restrictions, subgroups with insufficient sample sizes were combined into a single OTH group to maximize statistical power.

PS performance was evaluated using the area under the receiver operating characteristic curve (AUC), where all individuals without type 1 diabetes were classified as controls. The AUC was calculated for all participants and for individual ancestry groups within each biobank. Differences in AUC (ΔAUC) between scores were assessed using DeLong’s test.

We selected the PS with the highest performance and conducted a random-effects meta-analysis stratified by ancestry across four biobanks: AoU, Genomic Health Initiative at Endeavor Health (GHI), Penn Medicine BioBank (PMBB), and Geisinger MyCode (MyCode). MGBB was excluded from the meta-analysis because it was used in the development of T1D MAPS.

To assess the relationship between T1D MAPS and type 1 diabetes risk, we divided each biobank cohort into twenty bins, where each bin represents 5% of T1D MAPS values among individuals with type 1 diabetes in MGB Biobank. Odds ratios were computed in each biobank and for each T1D MAPS bin by comparing individuals within that interval to the entire cohort, for both screening (type 1 diabetes vs. controls) and diagnostic purposes (type 1 diabetes vs. type 2 diabetes). Lifetime risk for type 1 diabetes was also computed for individuals with T1D MAPS scores above each 5^th^ percentile, assuming a population prevalence of 0.3%. Study-specific odds ratios and lifetime risk estimates were then meta-analyzed across biobanks. Meta-analyses were performed using a fixed effect meta-analysis or random-effects meta-analysis with restricted maximum likelihood estimation of τ^2^ and Hartung-Knapp confidence intervals based on between-cohort variability. All analyses were conducted in R using pROC (v1.19.0.1) and meta (v8.2.1).

### Biobank Cohorts

- **Mass General Brigham Biobank (MGBB)**: MGBB is a repository linked to electronic medical records at the MGB Hospital system in Boston, Massachusetts [11]. Data from MGBB were current as of October 2022. Individuals were genotyped using the Illumina Multi-Ethnic Genotyping Array or the Illumina Infinium Global Screening Array [13]. Genome-wide imputation was performed using the Trans-Omics for Precision Medicine (TOPMed) r2 reference panel [14]. Imputation of HLA variants was performed with HLA-TAPAS [15,16]. Type 1 diabetes was defined based on manual review of medical records by a trained medical reviewer, as described previously [17]. Individuals needed to meet all of the following criteria: type 1 diabetes diagnosis confirmed by endocrinologist or primary care physician; current use of basal/bolus insulin regimen or insulin pump; and no secondary cause of diabetes listed in the medical record.
- **All of Us (AoU):** The All of Us research program is a longitudinal cohort study across the U.S. with detailed survey data and health information [10]. We performed analyses with the All of Us Controlled Tier Dataset v8 release. Genetic data were obtained from short-read whole-genome sequencing [18]. HLA variants were assembled from whole-genome sequencing data using Kourami [19]. Type 1 diabetes cases were identified using the eMERGE phenotyping algorithm [20], and all other individuals were classified as controls. Briefly, cases had a type 1 diabetes diagnosis code, were prescribed insulin, had no other glucose-lowering medications, and had no secondary causes of diabetes (e.g. drug-induced).
- **Genomic Health Initiative (GHI):** The Genomic Health Initiative (GHI) is the institutional genetic research biobank at Endeavor Health, a healthcare system in the Chicagoland area [21]. GHI participants provided informed consent, including permitting access to electronic health records (EHRs) for research, which was used to establish the type 1 diabetes (T1D) phenotype [22]. T1D cases were classified via their EHRs having the following International Classification of Diseases (ICD) codes: E10 (ICD10) or 250.xx ending in odd (ICD9). Germline DNA from blood samples was sequenced using low-coverage whole genome sequencing (lcWGS). Variant imputation was performed using a previously described method based on 1,000 Genomes Phase 3 haplotype reference panel [23]. Imputed SNPs were excluded from further analyses if there was (1.) posterior probability of <0.90, (2.) Hardy–Weinberg equilibrium deviation P < 10^−6,^ or (3.) allele frequency difference >0.05 compared with 1,000 Genomes Phase 3 reference panel.
- **Penn Medicine BioBank (PMBB):** PMBB collects genetic and electronic health record data from consenting individuals throughout south-central Pennsylvania, New Jersey, and northern Delaware as part of the University of Pennsylvania Health System [24]. Participants were genotyped using either the Illumina Infinium Global Screening Array or Genotyped by Sequencing capture. Imputation and phasing were performed on the Michigan TOPMed Imputaton Server using Minimac v4.1.6, eagle v2.4, and the TOPMed r3 reference panel. HLA imputation was also performed on the Michigan Imputation Server, using the HLA imputation 2.0.6 pipeline with the Four-digit Multi-ethnic HLA reference panel v2. Type 1 diabetes cases were identified using the eMERGE phenotyping algorithm [20].
- **Geisinger MyCode (MyCode):** Geisinger is an integrated health system serving predominantly rural populations across north-central and northeastern Pennsylvania through a network of more than 70 inpatient and outpatient facilities. The Geisinger MyCode Community Health Initiative is a longitudinal cohort that consent participants who contribute biospecimens—including blood, serum, and DNA—for research. These biospecimens are linked to participants’ longitudinal EHR data since 1996, enabling integrative studies of genetics, lifestyle, environmental exposures, and health outcomes [25]. Samples (175,477) were genotyped using Infinium OmniExpress Exome array (Illumina), and Infinium Global Screening Arrays (GSA-24v1.0 and GSA-24v2.0, Illumina). Genotypes for each array were imputed to the TOPMED reference panel (97,256 deeply sequenced genomes) with a GRCh38 build using the TOPMED Imputation Server. Imputation of HLA variants was performed with HLA-TAPAS pipeline (16). Type 1 diabetes was defined using an algorithm based on diagnosis codes, medication prescriptions, C-peptide levels, and/or pancreatic autoantibodies [26].

### Data And Resource Availability

Data from the All of Us Research Program are available to authorized users on the All of Us Researcher Workbench. Data from other biobanks are available to researchers affiliated with the respective institutions. The code used to generate polygenic scores are available at the following links:

T1D GRS2: https://github.com/sethsh7/PRSedm

T1D GRS2’: https://github.com/sethsh7/PRSedm

T1D GRS_HLA_: https://github.com/damichalek/T1DGC_GRS_HLA

T1GRS: https://github.com/Gaulton-Lab/t1grs

TA-PS: https://zenodo.org/records/15632544

T1D MAPS: https://github.com/snam-mgh/T1D_MAPS

## Results

Across the five biobanks, we analyzed 2,782 individuals with type 1 diabetes (2,132 EUR, 650 non-EUR) and 546,577 controls (340,536 EUR, 206,041 non-EUR) (Supplementary Table 1).

First, we compared the performance of each PS in AoU. The distributions of each PS differed between controls and type 1 diabetes cases within each ancestry group (Fig. 1). For each score, we selected the 90th percentile among controls with EUR ancestry in AoU as a fixed threshold to discriminate between cases and controls. Using this approach, the scores showed variable performance across ancestries. Overall, lower sensitivity was observed in non-EUR populations, while specificity was generally preserved. T1D GRS2 showed particularly low sensitivity in AFR ancestry (5.7%) despite high specificity (98.9%), whereas there was a more balanced profile in EUR ancestry (specificity 90.0%, sensitivity 65.9%). Compared to T1D GRS2, T1D MAPS showed improved sensitivity overall (56.7% vs. 51.2%), but specificity declined in several non-EUR groups. Comparable patterns were observed for T1D GRS_HLA_ and TA-PS. These findings show that using a European-derived threshold can reduce sensitivity in non-EUR populations, particularly in AFR ancestry.

**Figure 1.**
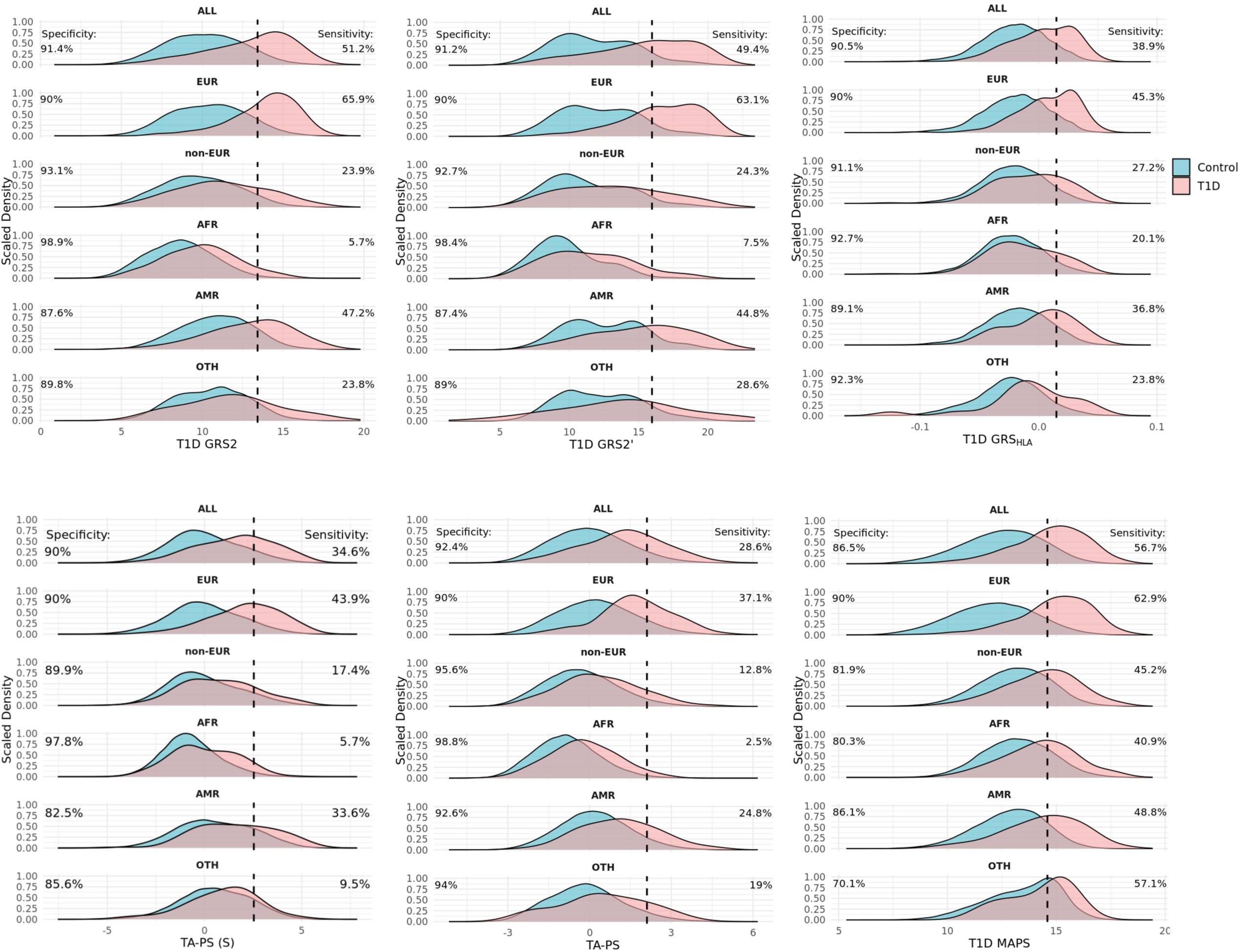
Density Plots of Polygenic Scores Across Ancestries in AoU. Distribution of T1D GRS2, T1D GRS2’, T1D GRS_HLA_, TA-PS (S), TA-PS, and T1D MAPS scores in control (blue) and type 1 diabetes (T1D; pink) groups across ancestries in the AoU cohort. Densities were scaled by their maximum value so that the y-axis ranges from 0 to 1. Vertical dashed lines represent the 90th percentile value among controls with European ancestry in AoU. Specificity and sensitivity percentages are shown for all participants (ALL) or for each ancestry group, including European (EUR), non-European (non-EUR), African (AFR), Admixed American (AMR), and Other (OTH).

To evaluate the relative performance of these scores, we calculated the AUC of each PS and assessed the difference in AUC compared to T1D GRS2, currently the most widely used type 1 diabetes PS. In AoU, T1D MAPS achieved the highest overall performance across ancestries (AUC_ALL_=0.801), followed by T1D GRS2 (AUC_ALL_=0.784) (Supplementary Table 2, Fig. 2a). Among EUR individuals, T1D MAPS and T1D GRS2 showed similar performance (ΔAUC_EUR_=−0.007, *p*=0.17). Other scores had significantly lower performance compared to T1D GRS2. Among non-EUR individuals, T1D MAPS significantly outperformed T1D GRS2 (ΔAUC_non-EUR_=0.052, *p*=3.8×10^−4^). No other score had a significantly higher AUC than T1D GRS2 in non-EUR populations, and this was consistent across AFR and AMR subgroups.

**Figure 2.**
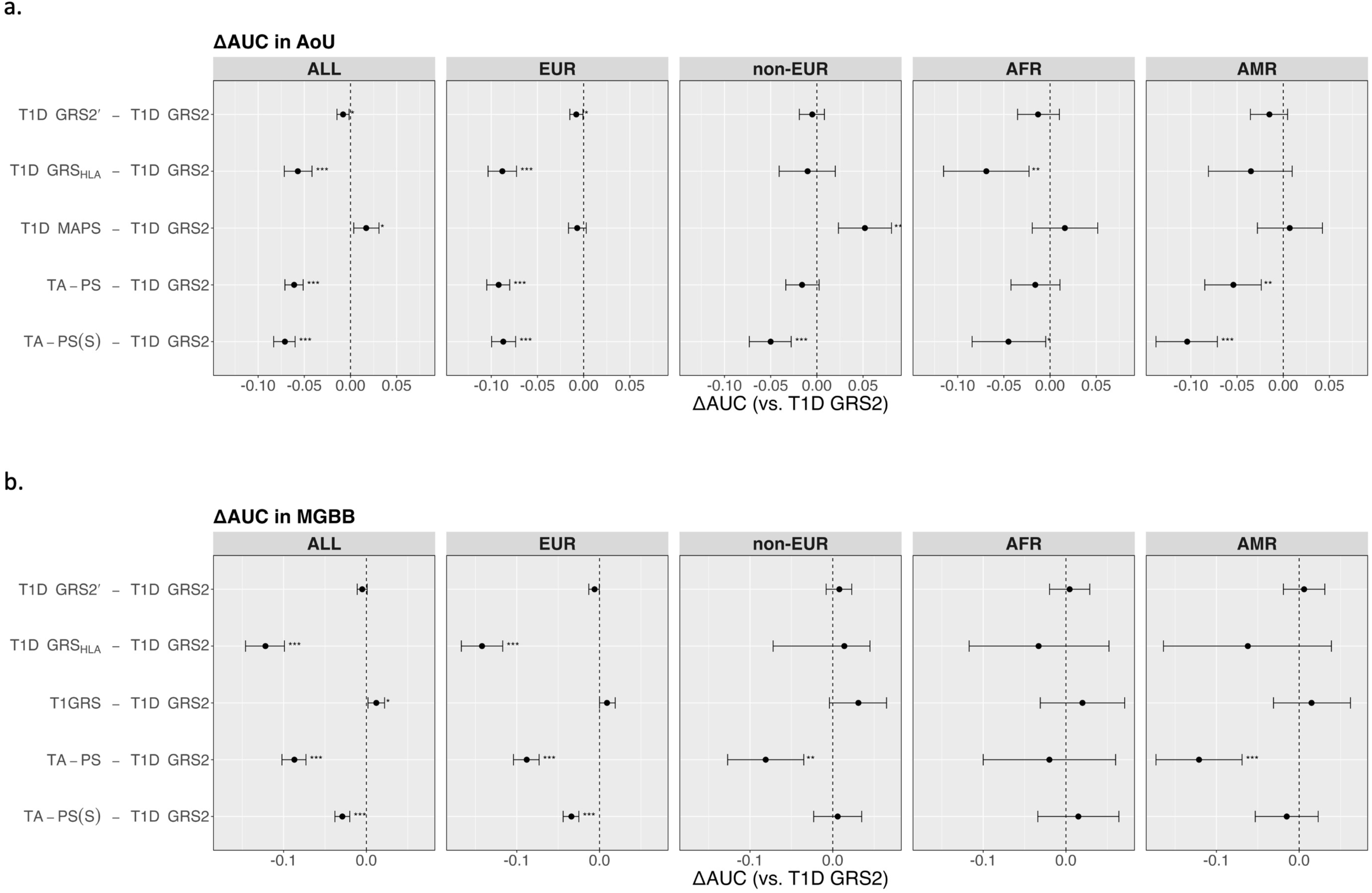
Polygenic Score Performance Across Ancestries in AoU and MGBB. Comparison of ΔAUC values (with 95% confidence intervals) relative to T1D GRS2 for various polygenic scores – including T1D GRS2’, T1D GRS_HLA_, T1GRS, TA-PS, TA-PS (S), and T1D MAPS – in the All of Us (a) and MGBB (b) cohorts. Scores were evaluated for all participants (ALL) or for each ancestry group, including European (EUR), non-European (non-EUR), African (AFR), or Admixed American (AMR). Vertical dashed lines represent no change in AUC. Statistical significance is indicated by p < 0.05 (*), p < 0.01 (**), and p < 0.0001 (***).

We were unable to evaluate T1GRS in AoU, as privacy restrictions prevented us from imputing HLA variants using the Michigan Imputation Server. However, we evaluated T1GRS along with other PS in MGBB. T1GRS showed a small but statistically significant improvement across all ancestry groups (ΔAUC_ALL_=0.012, *p*=0.014). Among non-EUR individuals, there was no significant difference between T1GRS and T1D GRS2 (ΔAUC_non-EUR_=0.031, *p*=0.081). No other score significantly outperformed T1D GRS2 overall or within ancestry-specific subgroups (Fig. 2b). Given that MGBB was used for the development of T1D MAPS, this score was not evaluated in MGBB to avoid potential overfitting.

Since T1D MAPS showed the highest performance in AoU, we next conducted a meta-analysis comparing ΔAUC between T1D MAPS and the widely used T1D GRS2 across four biobanks, excluding MGBB. In the EUR cohort (Fig. 3a), the pooled meta-analysis estimate showed no statistically significant difference between T1D MAPS and T1D GRS2 (ΔAUC_meta EUR_=0.016, 95% CI [−0.02, 0.05], *p*=0.33). In the non-EUR group, the pooled meta-analysis showed a statistically significant improvement for T1D MAPS over T1D GRS2 (ΔAUC_meta non-EUR_=0.049, 95% CI [0.03, 0.07], *p*=1.7×10^−7^), demonstrating a reproducible increase in discrimination across non-EUR populations (Fig. 3b).

**Figure 3:**
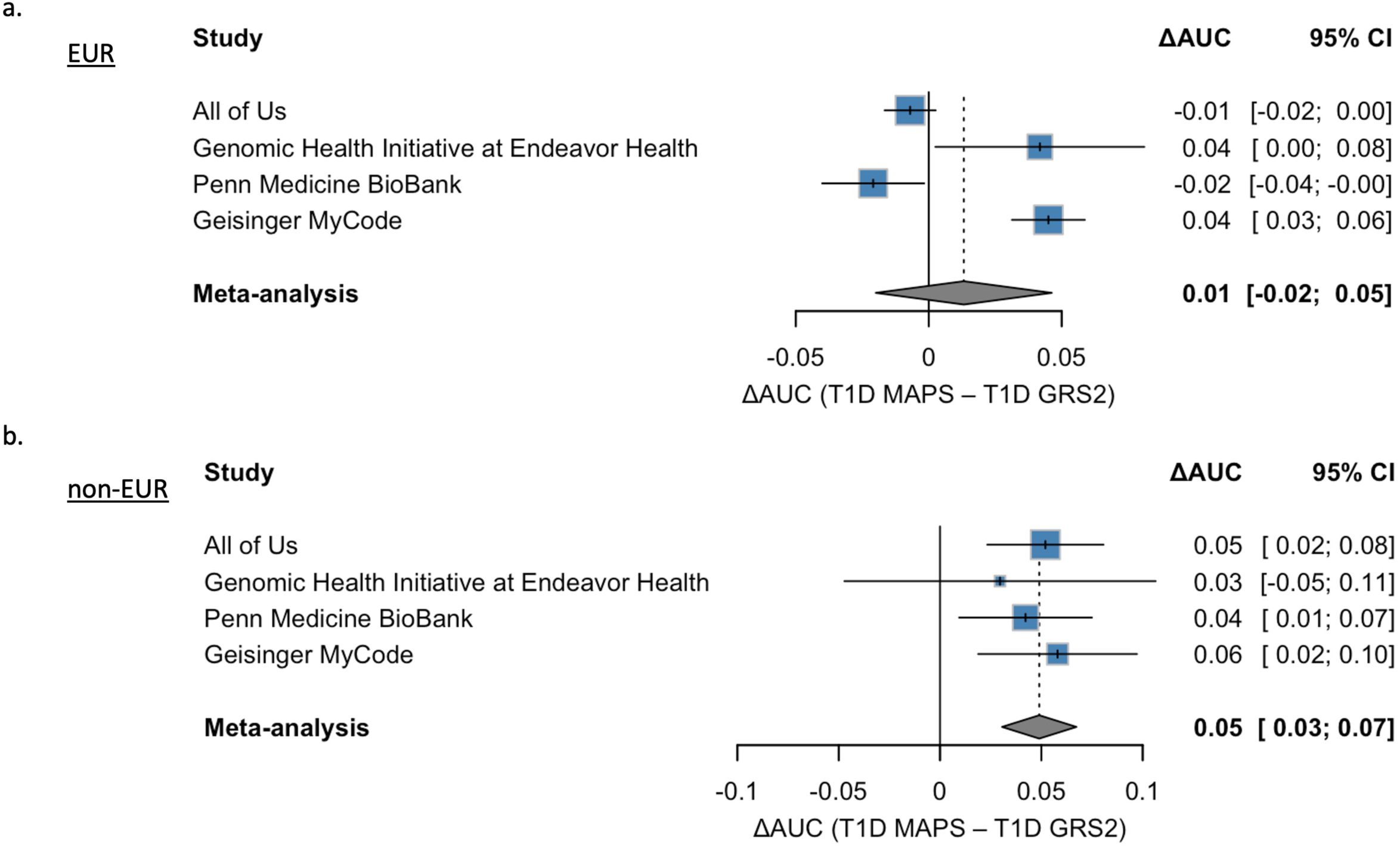
Meta-analysis of T1D MAPS vs. T1D GRS2. Meta-analysis of ΔAUC values (with 95% confidence intervals) comparing T1D MAPS to T1D GRS2 across four cohorts (All of Us, Genomic Health Initiative at Endeavor Health, Penn Medicine BioBank, and Gesinger MyCode), stratified by European (EUR; a) and non-European (non-EUR; b) ancestries. Blue squares indicate individual cohort ΔAUC estimates, while the diamond represents the overall random-effects meta-analysis summary (EUR: ΔAUC = 0.013, *p* = 0.44; non-EUR: ΔAUC = 0.049, *p* = 1.7 × 10^−7^).

We next performed a meta-analysis of study-specific odds ratios to generate clinically-actionable risk estimates for type 1 diabetes using T1D MAPS (Figure 4). Initially, we meta-analyzed four biobanks (excluding MGBB). However, risk estimates in PMBB differed significantly from the pooled estimates of the remaining cohorts (whole-curve tests for each outcome, *p*<0.001, Supplemental Table 3), with odds ratios and lifetime risk estimates approximately half of those observed in the other cohorts. Therefore, PMBB was excluded from the disease risk meta-analyses. This substantially reduced between-cohort heterogeneity for screening (I^2^: 60.4% vs. 25.8%) and diagnostic (I^2^: 62.7% vs. 25.3%) odds ratio analyses. Furthermore, the proportion of bins with significant heterogeneity (*p*<0.05) fell from 65% to 10% (screening odds ratio) and to 15% (diagnostic odds ratio) after excluding PMBB. Because the remaining between-cohort heterogeneity was low, a fixed effect meta-analysis was performed and demonstrated that T1D MAPS scores in the top decile of the type 1 diabetes distribution had a 24.6-fold odds of type 1 diabetes compared to controls (Supplemental Table 4) and 23.6-fold odds of type 1 diabetes compared to type 2 diabetes (Supplemental Table 5).

**Figure 4.**
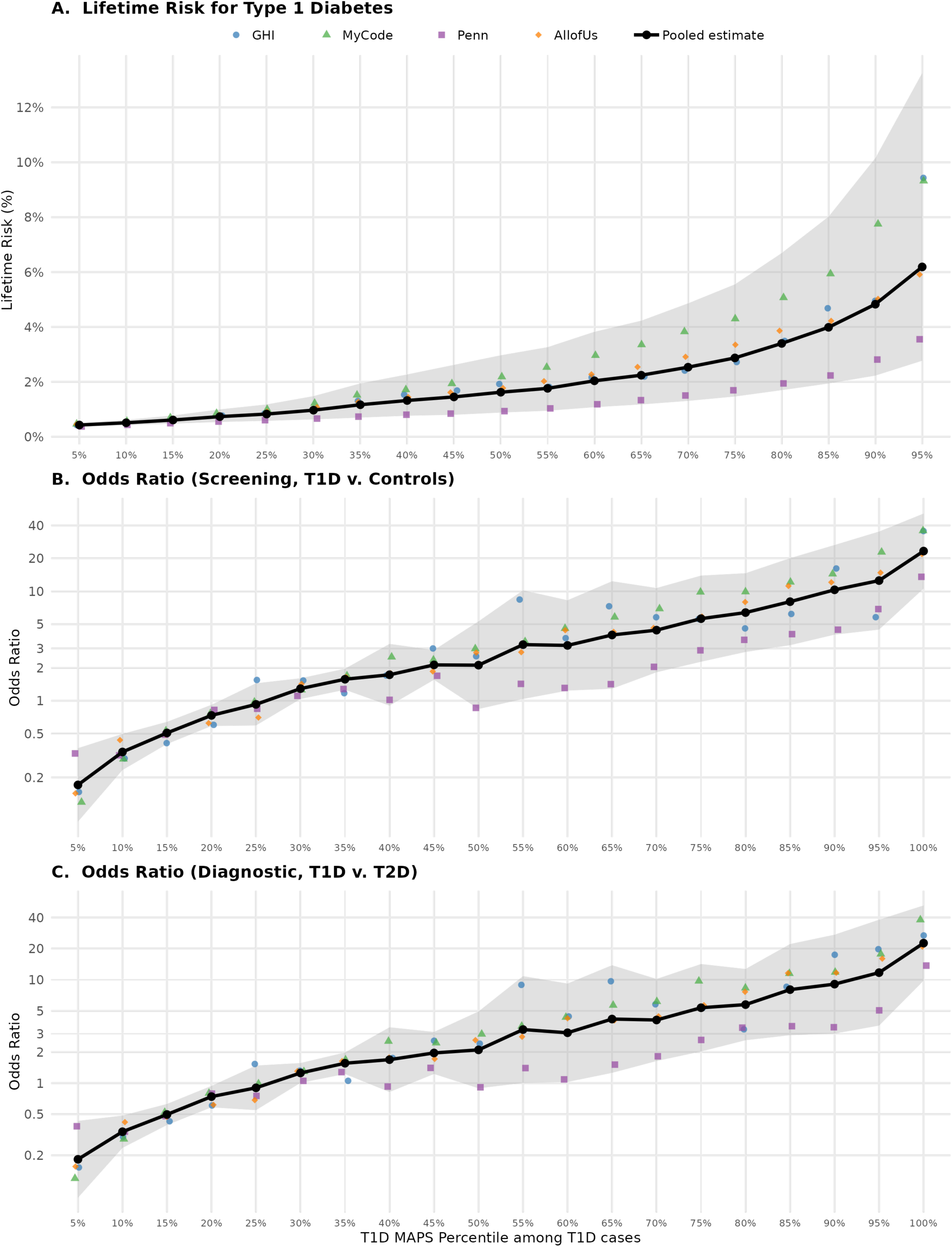
Lifetime Risk and Odds Ratio for Type 1 Diabetes at Different Values of T1D MAPS. (a) Lifetime Risk for type 1 diabetes (assuming population prevalence of 0.3%), (b) Odds Ratio for type 1 diabetes for screening (T1D vs. controls), and (c) Odds Ratio for type 1 diabetes for diagnostic purposes (T1D vs. T2D) in Genomic Health Initiative at Endeavor Health (GHI, blue), Gesinger MyCode (green), Penn Medicine BioBank (Penn, purple), All of Us (orange), and meta-analyzed pooled estimate (black) at different values of T1D MAPS. T1D MAPS percentiles were calculated among those with type 1 diabetes in MGB Biobank, and the same raw score thresholds were applied to each biobank. For odds ratios, the entire biobank was used as the reference population. The shaded region represents the 95% confidence interval for the pooled estimate.

We next analyzed lifetime risk for type 1 diabetes at various T1D MAPS thresholds. A random-effect meta-analysis with restricted maximum likelihood estimation of τ^2^ and Hartung-Knapp confidence intervals was used given that heterogeneity for lifetime risk remained high, even after excluding PMBB (I^2^: 97.5% vs. 85.0%). Individuals with T1D MAPS scores in the top decile of the type 1 diabetes distribution had greater than 6.0% lifetime risk of type 1 diabetes (Supplemental Table 6), rising further in the top 5% of the distribution. This T1D MAPS threshold (and even lower scores) are clinically actionable, with lifetime risk similar to or higher than that seen in first-degree relatives of individuals with type 1 diabetes [27].

## Discussion

In this multi-biobank evaluation of several type 1 diabetes PS, T1D MAPS consistently showed the highest performance. Notably, after applying a fixed score threshold to classify cases and controls, T1D MAPS displayed reasonably high sensitivity and specificity across ancestry groups, potentially avoiding the need for population-specific risk thresholds [28]. This has important clinical implications, as a single threshold could be applied across diverse patient populations without ancestry-specific calibration, enabling more consistent and scalable use of genetic risk scores in screening and risk stratification.

In individuals of EUR ancestry, T1D MAPS and T1D GRS2 demonstrated comparable performance. However, T1D MAPS significantly outperformed T1D GRS2 in non-EUR populations, with a pooled ΔAUC of 0.052 in a meta-analysis across four biobanks. This improvement is meaningful in the context of high-performing genetic predictors and may have important implications for equitable risk stratification. Furthermore, the consistent performance of T1D MAPS across independent biobanks underscores its robustness. Differences in recruitment approaches, genotyping platforms, and healthcare systems did not materially alter the direction of effect, though it did impact the magnitude of effect.

PS portability remains a major challenge in understanding human genetics. We observed higher predictive performance in EUR compared to non-EUR populations. Importantly, however, the relative improvement of T1D MAPS over T1D GRS2 in non-EUR populations suggests that model construction strategies can partially mitigate ancestry-related performance decay. Our results indicate T1D MAPS may improve discrimination in non-EUR populations without losing performance in EUR populations. Furthermore, T1D MAPS conveys clinically actionable effect sizes across populations. Using broader variant sets, better weighting, or multi-ancestry training data may enhance cross-population generalizability. While previous studies have demonstrated that clinical use of T1D PS may improve diagnostic accuracy [29], clinical implementation was hindered by differential score performance across ancestry groups. T1D MAPS now provides a T1D PS that might be more equitably implemented with less variability in interpretation by genetic ancestry. These findings highlight the need to include diverse populations in GWAS and PS development.

There are several limitations to consider. First, because of privacy restrictions and small sample sizes, we could not analyze all ancestry subgroups separately. To maximize power, some non-EUR groups were combined, which may obscure ancestry-specific effects. Additionally, the number of type 1 diabetes cases in some non-EUR strata was limited, reducing precision within individual cohorts. Larger studies with better representation of specific ancestry groups are needed for more granular comparisons.

Second, we observed lower AUC values compared to previously published results for certain polygenic scores, such as T1D GRS2. Performance estimates depend on the accuracy of the type 1 diabetes case-control phenotyping algorithm and the completeness of the underlying clinical and health records data. Misclassification or differences in data collection across biobanks may have influenced AUC values, odds ratios, and type 1 diabetes lifetime risk between cohorts. Importantly, the calculated odds ratio and lifetime risk are likely underestimates, as adult biobanks may miss adult-onset type 1 diabetes. Likewise, age at diabetes diagnosis can influence score performance. Individuals diagnosed with type 1 diabetes earlier in life may carry higher genetic risk. Consequently, PS developed in early-onset cohorts may have decreased performance when applied to more heterogeneous cohorts that include later-onset cases.

Finally, implementation challenges limited our ability to evaluate T1GRS. In AoU, because we were unable to access the Michigan Imputation Server, nine HLA variants could not be mapped and we were unable to compute T1GRS. In MGBB, we successfully used the Michigan Imputation Server, but two HLA-region variants were missing. Given the importance of the HLA region in type 1 diabetes risk, omitting these variants may have attenuated performance and highlights the challenges in harmonizing and implementing PS across biobanks. Although we were able to implement a near-complete version of T1GRS in MGBB, the greater degree of missing HLA information in AoU precluded a direct comparison of T1GRS and T1D MAPS within the same cohort. Future studies in cohorts with fully harmonized HLA data will be important to allow for direct comparisons of these approaches and to better understand their relative performance. Notably, however, among non-EUR individuals in MGBB, there was no significant difference between T1GRS and T1D GRS2, which were both trained using individuals with EUR ancestry.

In conclusion, T1D MAPS demonstrated a consistent and statistically significant improvement over T1D GRS2 in non-EUR populations, while showing comparable performance in EUR individuals. These findings underscore the importance of developing and evaluating genetic risk models in multi-ancestry populations to ensure equitable predictive performance. Furthermore, challenges in implementing T1GRS highlight the necessity of developing PS that are portable across different genotyping or sequencing platforms. Although we assessed several established PS, emerging multi-ancestry and machine learning-based approaches may offer additional gains and warrant direct comparison in future studies. Continued expansion of diverse cohorts and refinement of cross-ancestry modeling strategies will be critical to improving the accuracy, generalizability, and clinical utility of type 1 diabetes genetic risk prediction.

## Data Availability

All data produced in the present study are available upon reasonable request to the authors

## Personal Thanks

We gratefully acknowledge All of Us participants for their contributions, without whom this research would not have been possible. We also thank the National Institutes of Health’s All of Us Research Program for making available the participant data examined in this study. We are also grateful to the participants of the Geisinger MyCode Community Initiative who consented to the use of their anonymized genetic and clinical data for research. We thank the Geisinger-Regeneron DiscovEHR collaboration for making the genotype and phenotype data available for research.

## Funding

SN and AJD are supported by National Institutes of Health (NIH) / National Institute of Diabetes and Digestive and Kidney Diseases (NIDDK) K23 DK140643. RJK is supported by NIDDK K12DK133995. MSU, AKM, and JMM are supported by U01HG011723.

## Conflicts of Interest

MSU and JMM are involved in a research collaboration with Novo Nordisk. MSU has consulted for Novo Nordisk. LKB has received consulting honoraria from Novo Nordisk, Eli Lilly, Endogenex, Sanofi, Dexcom, Bayer, Xeris, Amgen, and Pfizer. JCF has received consulting honoraria from Alveus Therapeutics and is a PI on different research projects funded by Novo Nordisk. SN, RJK, JL, RM, HT, KT, MV, AH, JW, AJM, ARS, JX, BP, DJC, ULM, JCF, AKM, and AJD have nothing to disclose.

## Author Contributions and Guarantor Statement

SN, JCF, MSU, and AJD designed the study. SN implemented the polygenic scores, performed the meta-analysis, and wrote the first draft of the manuscript. RJK calculated the odds ratios and disease risk at different values of T1D MAPS. KT, MV, AH-C, AKM, and JMM analyzed data in All of Us. JL, DJC, and ULM analyzed data in Geisinger MyCode. RM and BP analyzed data in Penn Medicine BioBank. HT, JW, AJM, ARS, JX, and LKB analyzed data in the Genomic Health Initiative at Endeavor Health. All authors approved the final version of the manuscript. AJD is the guarantor of this work and, as such, had full access to all the data in the study and takes responsibility for the integrity of the data and the accuracy of the data analysis.

## Prior Presentation

Parts of this study were presented in abstract form at the 86th Scientific Sessions of the American Diabetes Association, New Orleans, LA, 5-8 June 2026.

## Notes

### Author Declarations

Analysis of Mass General Brigham Biobank was approved by the Mass General Brigham Institutional Review Board (study protocol 2016P001018). Analysis of the All of Us cohort was approved by an institutional Data Use and Registration Agreement between Mass General Brigham and the All of Us Research Program (study protocol 2020P002213).

